# Community healthcare workers’ experiences during and after COVID-19 lockdown: a qualitative study from Aotearoa New Zealand

**DOI:** 10.1101/2021.11.26.21266650

**Authors:** Eleanor Holroyd, Nicholas J. Long, Nayantara Sheoran Appleton, Sharyn Graham Davies, Antje Deckert, Edmond Fehoko, Megan Laws, Nelly Martin-Anatias, Nikita Simpson, Rogena Sterling, Susanna Trnka, Laumua Tunufa’i

**Affiliations:** School of Clinical Sciences, Auckland University of Technology, Auckland, Aotearoa New Zealand; Department of Anthropology, London School of Economics and Political Science, London, United Kingdom; Centre for Science in Society, Victoria University of Wellington, Aotearoa New Zealand; School of Languages, Literatures, Cultures, and Linguistics, Monash University, Melbourne, Australia, and School of Social Sciences and Public Policy, Auckland University of Technology, Auckland, Aotearoa New Zealand; School of Social Sciences and Public Policy, Auckland University of Technology, Auckland, Aotearoa New Zealand; School of Māori Studies and Pacific Studies, University of Auckland, Auckland, Aotearoa New Zealand; University of Waikato, University of Waikato, Hamilton, Aotearoa New Zealand; School of Social Sciences, University of Auckland, Auckland, Aotearoa New Zealand

**Keywords:** Community Nursing, Personal Care Assistants, Pandemic, Feelings of Duty, Fear of Contagion, Work-Life Balance Issues

## Abstract

Shortly after the COVID-19 pandemic reached Aotearoa New Zealand, a stringent lockdown lasting seven weeks was introduced to manage community spread of the virus. This paper reports the findings of a qualitative study examining how lockdown policies impacted upon the lives of those caring for community-based patients. The study involved nationwide surveys and ethnographic interviews with 15 registered nurses (RN) employed in community settings, two community midwives, and five personal care assistants (PCAs).

During the strict lockdown levels 4 and 3, RNs and PCAs in the community showed considerable courage in answering their “call to duty” by taking on heightened care responsibilities and going “the extra mile” to help others. They faced significant risks to personal and professional relationships when they were required to take on additional and complex responsibilities for community-based patients. Despite, and sometimes due to the hypervigilant monitoring of their personal protective equipment (PPE), the need to safeguard family and community members generated considerable stress and anxiety. Many also faced personal isolation and loneliness as a result of lockdown restrictions. Although ‘care’ and ‘kindness’ became social expectations throughout Aotearoa New Zealand during the lockdown, RNs and PCAs who were already doing care work in patient homes had to do more.

This article makes five core service delivery and policy recommendations for supporting community-based nurses and PCAs in respiratory disease pandemics: acknowledging the crucial role played by community-based carers and the associated stress and anxiety endured, through championing respect and compassion; demystifying the “heroism” or “self-sacrifice” projected onto care workers to facilitate boundary setting; the timely provision of adequate protective equipment; improving remuneration with adequate provision for time off; and regular counselling, peer support groups, and education on work-life balance delivered by support workers in recognition of stressors arising from these complex and isolated working conditions.

**What is known about the topic:**

- Nurses and personal care assistants play a pivotal role in community responses to pandemics.
- The COVID-19 pandemic has intensified many community healthcare workers’ clinical duties.
- Pandemics pose risks to healthcare workers’ physical and mental wellbeing.

**What this paper adds:**

- Community healthcare workers pressured themselves to be a “good carer” or “hero” during the lockdown.
- Caring *for* patients in the community also became about caring *about* patients, further intensifying workload.
- The COVID-19 pandemic has negatively impacted community healthcare workers’ relationships, as well as their wellbeing. Impacts continued even once the virus was eliminated.
- Need for recognition of this workforce distinct from other care workers.

## Introduction

Nurses and PCAs are the largest groups of front-line healthcare service workers delivering care in communities in Aotearoa New Zealand (hereafter Aotearoa), yet they are often the least recognized group of healthcare providers (Green, 2020; Lukewich et al., 2021). These workers provide a spectrum of services ranging from supporting daily living to administering complex pharmaceutical interventions. They are pivotal in ensuring effective and ethical care provision for “home-based” patients during pandemics (Ives et al., 2009; World Health Organization, 2020). Community based registered nurses (RNs) and personal care assistants (PCAs) deliver individualised patient care, most often in patient’s homes, away from their colleagues and institutional support systems. They often work irregular shifts, in close physical contact with patients, and in community settings, putting them at heightened risk of infection.

The impact of COVID-19 on healthcare workers, in particular RNs, has been widely noted globally, with commonly reported difficulties including anxiety and stigma as a result of potential workplace exposure to COVID-19, concerns about inadequate PPE provision, exhaustion from proliferating workloads, and the trauma of having patients die of COVID-19 (Bagnasco et al., 2020: Maben & Bridges, 2020; Nyatanga, 2021). Such issues can contribute to burnout and attrition from the workforce (Fernandez et al., 2020). Nevertheless, as De los Santos and Labrague (2021, p54) note, the literature has often placed more emphasis on hospital workers than staff working in the community. Hospital staff, particularly those treating COVID-19 inpatients, certainly face immense challenges - and surveys from Israel indicate that they have experienced a bigger drop in occupational satisfaction during the pandemic than community nurses (Savitsky et al., 2021). However, those providing healthcare in the community face distinct challenges that warrant consideration in their own right.

With patients unable to access regular clinics due to lockdowns, community healthcare providers have had to adopt new protocols, ranging from teleconsulting to providing clinical services in patients’ homes (Corcoran, 2021; Oldman, 2020; Penfold, 2020; Stanley & Rawlinson, 2021; Yi et al., 2020). Mastering these new procedures can itself be a source of stress (Maben & Bridges, 2020). Moreover, although some healthcare systems implemented triage systems to reduce pressures on staff (Yi et al., 2020), many community nurses have seen their workloads intensify given the increased demand for home-care (Bowers et al., 2021; Green et al., 2020), and the medical complexities of treating and supporting recovery from COVID-19 (Green, 2021; Sun et al., 2021; Trueland, 2020). Globally, community healthcare workers have also been tasked with enforcing social distancing between patients and their families (Sun et al., 2020).

To date there has been little research on the experience of community healthcare workers in Aotearoa – a country that has had a distinctive experience of the pandemic, given the rapid elimination of COVID-19 (Cousins 2020; Kvalsvig & Baker, 2021). Elimination followed the swift imposition of stringent lockdown measures restricting most people’s personal movement to a single ‘bubble’: an exclusive social network, usually coterminous with a single household (Long et al., 2020). Community healthcare workers were seen by healthcare management as integral to the lockdown’s success: they were instructed to be public health ‘role models’ and ‘educators’ (Brinkman, 2020) and have since been celebrated as ‘heroes’ (McClunie-Trust, 2021).

This paper draws on community healthcare workers’ own accounts to map the key themes that emerged as areas of concern for them during lockdown. It corroborates international research on primary care workforces, showing that heavy workloads together with a lack of PPE can cause considerable psychological stress (De Kock et al., 2021). It also reveals how the stringent lockdown measures implemented in Aotearoa placed additional workload pressures on this group of caregivers by rendering them compelled to “go the extra mile” and take on care duties extending beyond their formal remit. It further shows how the mental and relational strains of the pandemic have continued even since COVID-19’s elimination. It thereby highlights some of the negative corollaries of the elimination strategy in Aotearoa. These must be considered when planning future pandemic responses. We conclude with policy recommendations to help mitigate these issues.

## Methods

An exploratory approach guided the two phases of data collection, seeking to examine the experiences and challenges of delivering community healthcare under lockdown. Phase 1 involved analysing free-text answers from three non-probability online surveys, conducted at Levels 4, 3 and 2 of the lockdown (starting April 2020). These surveys were advertised via word-of-mouth and Facebook ad campaigns. We received a total of 3644 valid responses. Phase 2 involved an interpretative approach using one-off ethnographic interviews with 22 community-based health service workers which included 15 registered nurses (RNs) (public health, district, primary care, and mental health), two midwives, and five PCAs. Twenty of these participants were women and two were men. Interviews were conducted between May 2020 and August 2020.

### Sampling

Survey respondents were self-selected, with the nationwide Facebook ad campaign ensuring that the opportunity to participate was promoted as widely as possible. Interviewees were recruited from the health service workers who took part in the nationwide surveys, as well as via social media flyers and snowball networking. Nevertheless, and as is common for survey research in Aotearoa (Houkamau & Sibley, 2019) the sample skewed heavily towards women, Pākehā (New Zealanders of European descent), and university graduates.

In what follows we do not attempt to provide a statistically representative or exhaustive account of the ways nurses and PCAs experienced the lockdown in Aotearoa. By foregrounding themes and issues that were voiced by multiple respondents, we nevertheless aim to illuminate dynamics that we consider significant and widely shared.

### Approach to the Ethnographic Interviews

A guide for the interviews was developed from a literature review, as well as preliminary analysis of survey responses and aimed at asking:

1. What worked and what was challenging about your bubble and those of your patients?
2. What arrangements had been made to make the workplace and caregiving spaces to be COVID-secure?
3. What are your thoughts on these arrangements?
4. How did your household arrangements influence your work, care, and life under lockdown experience?

Interviews were conducted primarily by EH. They lasted between 20 and 40 minutes and were held on Zoom or by phone after digital written consent had been granted and participant confidentiality assured.

### Analysis

Audio recordings were transcribed verbatim by the interviewer or via Otter.ai. Analysis followed the reflexive approach of Braun and Clarke (2019). First steps included familiarisation with the data, with the transcripts read repeatedly, and generating initial codes that highlighted significant features of the data. These codes were then compiled into initial themes, and data was matched to each potential theme. The themes were then reviewed alongside the coded data and the complete data set, to provide a thematic map of our analysis. The themes were then refined and formally defined through weekly discussions among the research team over the course of several months. The finalized themes were then further defined and named. Transferability was achieved through drawing out thick descriptions to frame contexts, and settings (Liamputtong, 2019). Regular team discussion ensured confirmability, safeguarding against conclusions being unduly influenced by any one researcher’s bias or interests.

### Ethical considerations

Ethics approval for this research was granted by the Research Ethics Committee at the London School of Economics and Political Science (refs 11.08a; 11.08b; 11.08c) and the Auckland University of Technology Ethics Committee [ref 20/142]. It was further approved and ratified by the University of Auckland and Victoria University of Wellington’s Human Participants Ethics Committee. All names used are pseudonyms.

## Findings

Four main themes recurred across community health care workers’ accounts of living through and delivering care during the stringent lockdown in Aotearoa:

A. Going ‘the extra mile’ – Heightened professional responsibilities and additional care work associated with the lockdown,
B. The need to provide protection for self and family,
C. Strain on personal relationships,
D. Long-term ramifications of lockdown.

### (A) Going ‘The Extra Mile’ – Heightened Professional Responsibilities and Additional Care Work Associated with Lockdowns

The requirement that people other than essential workers stay within a single ‘bubble’ for seven weeks disrupted the flows of inter-household care that would usually support patients living in the community (Long, 2020). The deep relationships that many carers had developed with their patients prior to COVID-19 were thus heavily leaned upon during lockdown - a development which could take a toll on care workers at the coalface of community service delivery, who were left to absorb the risks. Moreover, many patients’ wellbeing was compromised by the seven-week period of extended confinement.

Respondents described how their patients - particularly those in rest homes - “got worse” or “deteriorated” under lockdown due to changes in their routine, such as not being allowed to see their families or go outside or being shut in their homes:

> […] it was like everything fell apart and those who had dementia got worse very quickly. So hard seeing family try and talk over walls and you could not really tell these patients what was happening.
>
> Pākehā RN.
>
> “[…] they picked up upon the stress of the outside world and it was like a spring just coiling up all the time.
>
> Pākehā RN.

Faced with such situations, participants overwhelmingly saw it as their duty to ‘go the extra mile’ and assist patients in need. A Māori PCA drew on their childhood and spoke about using whānau (family) principles to explain care and compassion they felt for their in-house patients.

> I would expect the same work of care for myself; I was brought up this way […] it is natural of me to care. So, I went the extra mile. I put laboratory test results in letterboxes and got extra groceries for those living alone.
>
> Māori PCA.

“Going the extra mile” often involved working longer hours:

> That is just what you do. You can’t switch off caring; it never goes away. What is wrong with going the extra mile? That is what my family did.
>
> Pākehā RN.

Increased workloads did not just arise from carers taking on more through personal moral obligations, but also because the number of safety protocols needed to mitigate risk or initiate protection increased dramatically during COVID-19. As one nurse articulated:

> It’s giving of yourself and then our workloads increased so much, with extra cleaning, tracing, and hygiene protocols these took soooo long. That’s why nursing is so exhaustive. It… it’s giving of yourself, then giving more at home, and in our family, and now along comes COVID-19.
>
> Pākehā RN

Despite their exhaustion, however, participants often emphasized how they “had to keep going” as they were driven by existing personal bonds with their patients. They took on extra personal tasks – which were often numerous and time-consuming, such as delivering groceries or running errands – without extra pay or time-off allowances. While they felt their extra work was valued by their patients and their patients’ families, they also experienced feelings of heroism and self-sacrifice from doing more than they were strictly required to as healthcare workers.

Apparent here is a moral notion of pandemic response, in which health service workers go above and beyond what is expected of them, and “good characters” emerge (Brazeau et al., 2010). Many PCAs expressed taking on an extended identity, adding new dimensions to their unique care relationship with vulnerable patients living in what were often very isolated conditions. The need to “keep on helping” was frequently invoked as an unquestioned moral imperative – that was “what nursing [or caring] was” and “what duty was”.

Moreover, most participants experienced a deepened connection to and with their increasingly vulnerable community-based patients. A very particular ethics of care was enacted in moments of “going the extra mile”; one RN outlined how “‘really caring” involved more than simply carrying out the “care work” that one was assigned:

> There’s a difference between really caring for a patient in the community and carrying out tasks. I am emotionally invested in these older people who have such tough lives and are scared of the virus, many do not see their families – they do not have anyone else in their bubbles. Caring is a feeling that I don’t think you can ever really explain as a nurse. With time, you gain connection, and you share part of yourself with them. Pākehā RN

Kelly et al. (2020) and Wright St Clair & Seedhouse (2005) have similarly shared narratives of patients experiencing times when caring went beyond just medical treatment and life support to include unanticipated acts that showed deep compassion and empathy, pointing to an existential difference between being cared “for” and being cared deeply “about” as a person.

Popular media depict carers, especially nurses, as dedicated and equipped with both the instinct and the motivation to serve. Further, health care workers are often imagined, and indeed expected, to be a self-sacrificing workforce that can be depended on beyond all odds (Shan et al., 2021). Many of our respondents had internalised such expectations and used them to evaluate themselves as carers and as moral persons; they could take satisfaction in having done what they thought was right, despite the increased workload that this created (see also Blanco-Donoso et al., 2021). Nurses and PCAs were not necessarily being *required* to do more but applying their own moral compass and extending themselves to do more for their patients, without any extra pay or formal recognition.

### (B) The Need to Provide Protection for Self and Family

Many interviewees described juggling complex work roles alongside protecting themselves and their families. Most of the PCAs interviewed were over 50 years of age and often shared households with people who were older and vulnerable, such as older spouses, elderly parents who had joined their bubble for lockdown, and/or immunocompromised family members. Moreover, even when respondents’ household members were not vulnerable, they often felt a deep imperative to protect them. A Pākehā PCA highlighted the complexity of such a situation:

> I am a home carer for older adults with severe health issues living alone. I needed to shower and dress them as well as feed some through tubes and give injections all while masked, plus [at least one other person was] living with me at home. So, to keep [them] safe I had three bubbles; I slept in a shed to isolate [them], so [they were] protected in [their] bubble, then I had all the work bubbles I needed to form with my home-based patients. I did this so my…so we would be alright and would not be contaminated. And on top of that people I know looked sideways in the supermarket at me.

To protect her family and herself from COVID-19, Sandra, a public health nurse, asked to undertake COVID-19 contact tracing duties (conducted over the computer and phone) to avoid going into patients’ homes as COVID-19 cases spiked. While Sandra was able to make this change, many others could not. For them, the emotional labour of community-based care work in a pandemic meant risking not only their patients’ health and wellbeing, but also their own, and that of those close to them (see also Holroyd & McNaught 2008; Major, 2008). While some devised strategies for managing the complex emotions this situation generated, others reported feeling overwhelmed. Tracey, a registered nurse, shared how she felt living with the fear of coronavirus each day: “I was so shaky all the time. I have to work as if nothing is happening, and I just want to be at home in my own bubble not here.”

This dual work of caring at work and home was made additionally stressful by the lack of access to adequate PPE. The nurses and PCAs spoke of feeling that they were at the “bottom of the pile” for the provision of equipment such as gowns, masks and gloves. A a Pākehā community nurse, said “I am working in total isolation with the physiotherapists all getting protective clothing ahead of me, despite the fact that they only saw patients for half an hour.” A community-based midwife had made gowns and masks in a sewing circle with other midwives (who could professionally share a bubble) so they all had enough PPE to attend home visits. Making their own PPE was considered not only to be protecting their patients, but also their family and community members.

### (C) Stresses on Personal Relationships

Although the lockdown in Aotearoa was, by international standards, relatively short-lived, it had far-reaching repercussions for many people’s relationships. Research participants often described how being confined to separate bubbles had attenuated once-close relationships. Kay, a PCA, described having drifted apart from her best friend, who lived on the other side of town and had become preoccupied with their own bubble. Whilst not unique to essential workers, such dynamics of estrangement often affected them acutely. As one Pākehā midwife explained, work was so stressful that many had “no mental or emotional energy left once off work” to interact with friends. These pressures were frequently compounded by the obligations they felt to provide continuing care and support to patients who they knew to be isolated, anxious, and/or at risk – for example, through phone calls in the evening and weekends to their patients in the community.

Both nurses and PCAs described facing community stigma as a result of their work. Some faced hostility and accusations of breaking lockdown rules when travelling for work, while others described being shunned or avoided in grocery stores by community members who feared they could be vectors of infection. Clara, a primary health care nurse described how one supermarket had refused her entry when she was dressed in her uniform. She also shared her sadness at having experienced stigma within some of her closest relationships. Some family had refused to see her, even several weeks after COVID-19 had been eliminated and Aotearoa had moved to Level 2. “They’re too afraid,” she explained, “they think I’m at higher risk of passing on COVID because of my job… and because of my job that meant my children were in daycare during level 3 too. They believed we posed a higher risk, despite these family members opening their bubble to other family”.

Proliferating workloads and anxieties regarding COVID-19 exposure could sometimes strain relationships within nurses’ and PCAs’ bubbles, a theme that many spoke about in their interviews and that they sought to address in a variety of ways. One midwife used creative writing to provide a meaningful connection with her partner, as well as renewing her relationship to God to seek strength during the lockdowns. This religious support was both spiritual and social, because while she was living in a work bubble in a home birth location she found support through her church community, especially their online poetry club. This indicates that while the lockdowns were particularly hard for in-home carers and nurses, some did find support through the tough times. Others, however, became worn down by needing to provide endless compassion and kindness and then go home and do it all over again – with nobody in particular caring for *them* as they continued to work without any guarantees of extra personal protective measures for themselves or their families.

### (D) Long-term ramifications of the pandemic and lockdown

The themes above have identified issues affecting the wellbeing of nurses and PCAs working in the community *during* the lockdown. However, although the most stringent restrictions on personal movement ‘only’ lasted seven weeks, the ramifications of the lockdown continue to be felt by many community healthcare workers.

In some cases, these ramifications reflect the ongoing impact of physical and, especially, mental health conditions that originated early in the pandemic. For example, even one year after the seven-week lockdown, several nurses felt that the heightened anxiety amongst their patients had not stopped, and that their experiences of tending to patients in isolation, coupled with their own need for constant vigilance, meant that they themselves now lived with fear of the unknown or threat of contagion. Some reported being unable to exercise control over this fear. For example, one Pākehā nurse described how it was: “very hard to calm down from there. I feel like a tap has been turned on, and all the anxiety of my life is running nonstop. I still don’t think it has stopped.”

Others indicated long-term impacts of the stigma they carried as suspected vectors of COVID-19, either because loved ones remained anxious that they might transmit the virus or because the damage to their relationships proved difficult to heal. In some cases, experiences of lockdown even led respondents to question whether they wanted to continue in their current roles. For Clara, the primary care nurse mentioned in the previous section, the lockdown had proven both eye-opening and disillusioning:

> I think it has reinforced how little nurses are valued in NZ, especially primary health care nurses. While supermarket workers got a temporary increase in pay to reflect the risk of working during this time, nurses got nothing. We got very little recognition and some people were rude to us… For many of us, it has left us feeling deflated and highly undervalued. It has definitely got me thinking of future career changes.

While the moral imperative to care for others had led to many research participants ‘going the extra mile’ in the acute moment of lockdown, the pride they took in ‘doing the right thing’ was offset not only by the acute stresses they felt at the time, but the lack of support and recognition they received from both the community and the government. Although the lockdown was relatively short-lived, its impact continues to be keenly felt.

## Discussion

This study reveals how important notions of ‘being a good carer’ and ‘doing the right thing’ were for community nurses and PCAs during the COVID-19 pandemic in Aotearoa. Their accounts present the pandemic as an extreme context requiring the repeated enactment of complex moral judgments. What is tested is the tensions between: nationally mandated health regulatory bodies’ ethical codes and practices (predicated upon the Hippocratic Oath of protection); rapidly reconfiguring what action can take place to do social good and avoid harm in situations of professional isolation and potential danger; and needing to protect loved ones. Navigating these tricky ‘competing responsibilities’ (Trnka & Trundle, 2017) contributed to the psychological pressures facing this often-overlooked workforce.

Determination to ‘do the right thing’, ‘go the extra mile’, and ensure they were ‘caring about’ their patients, rather than simply ‘caring for’ them, led many respondents to take on additional tasks outside their stated work remit. This added to workloads that were already soaring due to time-consuming COVID-19 protocols and increased demands for home care. Meanwhile, health care workers’ anxieties about the possibility of contracting or transmitting COVID-19 were compounded by moral anxieties about whether they truly were ‘doing the right thing’. Many felt inherently compromised: they were providing care and support to patients in the community who were profoundly isolated – and often deteriorating clinically – as a result of the lockdown. Of note is the heavily gendered need to absorb caring responsibilities inherent in the feminisation of caregiving with community carers going the extra distance both at home and at work. While the participants who responded to our call may have been the most likely to go above and beyond, hence not representative of *all* carers, the theme of ‘going the extra mile’ was sufficiently prominent to suggest it warrants close consideration in future pandemic planning.

By staying loyal to the call of duty and attempting to maintain order in the face of adversity, participants also perceived that they put their patients and their own families at risk. They showed considerable determination and courage; but they also felt exhausted and emotionally overwhelmed, sometimes displaying serious symptoms of mental distress. They often had to grapple with stigma, isolation and loneliness in their personal lives. Moreover, many felt poorly supported by existing organizational provisions – particularly regarding PPE.

The notion of an obligation to sacrifice without extra resources (PPE, occupational wellbeing support, etc.) has been used to portray nurses are model citizens, as if being framed as ‘heroes’ were a reward in itself, conferring status on previously invisible groups (Appleton et al., 2020). However, this in turn places extreme pressure on this workforce (Shan et al, 2021). While the literature observes that many healthcare professionals are willing to accept the risks of their occupation in a pandemic, others perceive the risks of their work to be too high (Brooks et al., 2018). This reluctance to step up may be especially so when their sacrifices are not adequately recognised and the additional support needed to continue in the job is denied. Such gaps in the institutional support of community-based healthcare workers have been documented to trigger adverse mental health amongst frontline care workers in private homes and domiciliary care agencies during COVID-19 (Nyashanu et al., 2021), and to impact upon retention (Halcomb et al., 2020; Sumner & Townsend-Rocchiccioli, 2003). As this study shows, it can also generate a sense of moral injury and injustice. When such actions were uncertain, delayed or given by their organizations to other health professionals first, it was perceived as an affront. In pandemic times, such failure to take action on the part of the institutions can compromise the interpersonal ‘good actions’ taking place.

To date, much literature on healthcare workers’ experiences of pandemics has focused on the clinical burdens, risks, and fears that arise as a result of COVID-19 transmission (e.g., Bagnasco et al., 2020; De los Santos & Labrague, 2021; Green, 2021; Yildirim et al., 2020). The case of Aotearoa, from which COVID-19 was rapidly eliminated following a highly stringent lockdown – is instructive for highlighting the additional pressures that can result as a result of pandemic control measures themselves. Restrictions on household mixing not only contributed to a deterioration in the wellbeing of community patients, but also deprived them of many forms of practical support, which nurses and PCAs then felt an added duty to provide. Meanwhile, the strict lockdown restrictions, in which one could not even meet outside for a socially distanced conversation or walk, heightened these workers’ own sense of loneliness and isolation. Opportunities to expand bubbles at Level 3 of lockdown could, in principle, have provided relief. In practice, however, few respondents took up this option, either because they faced stigma from prospective bubblemates or themselves worried that they might be vectors of infection (see also Long et al., 2020; Long et al., forthcoming; Trnka et al., 2021). Such findings highlight how social distancing – often framed as an ethical endeavour undertaken to ease pressure on health systems and healthcare workers (Jackson et al., 2020; Marchesi, 2020) – can actually generate distinct pressures for certain groups of the latter. Circumventing these pressures for this workforce (and other workforces sited at the interface of community and services) should be a priority for future pandemic planning. It is important that attention is provided to community care, personal resilience, and patient advocacy in nursing education programmes, together with adequate recompense for the critical ‘social infrastructures’ (Bear et al., 2021) that their care provides for vulnerable people and communities during pandemic times.

Lastly, we note a significant irony. In countries that have been continuously dealing with surges of COVID-19 cases since 2020, there has been a concentrated although somewhat belated effort to develop protocols that safeguard community-based carers (Ballard et al., 2020). However, because Aotearoa did so well in rapidly eliminating COVID-19, there has been limited recognition of the extreme hardships that certain groups, such as community nurses and PCAs, experienced during intense lockdown periods, nor policy developments aimed at addressing the professional and wellbeing issues that these caregivers are still experiencing as a result of the March-May 2020 lockdown. A final conclusion stemming from this study is thus that future pandemic planning should not only devise strategies for supporting community health workers through potentially protracted lockdowns, but must also think of ways to minimise and remedy the long-term repercussions of short-lived, but intensely stringent, modalities of infection control.

## Recommendations

Efforts to address the consequences of heightened community-based care responsibilities during a pandemic must start by acknowledging these as legitimate concerns and making a sincere commitment to reduce the associated stress and anxiety. The additional duties and risks taken on by community nurses and PCAs should be compensated via pay rises, especially in settings where other frontline workers receive such ‘rewards’ (as was the case for supermarket workers in Aotearoa). Moreover, staff must be treated with respect and compassion, and given adequate provision for time off. It is imperative that PPE is provided in a timely manner.

Regular counselling, peer support groups, and support in achieving a manageable work-life balance needs to be provided in recognition of stressors arising from these complex and isolated working conditions. These should ideally be provided by external support workers, although the literature suggests regular team meetings or ‘huddles’ can also prove beneficial (Franzosa et al., 2021). Support measures should extend well beyond the duration of the lockdown itself in recognition that the psychological and social effects of stringent pandemic control measures can be long-lasting. To these initiatives needs to be added the demystifying of the “heroism” or “self-sacrificing” placed on care workers to facilitate boundary-setting. Especially valuable during and after a pandemic, these are measures that should be considered best practice for supporting community healthcare workers at all times.

Additional support for isolated patients (such as social media befriending groups) should also be considered, to alleviate the burden on nurses and PCAs. Where the epidemiology permits, more flexible pandemic control measures (e.g., allowing people to meet at a physical distance outdoors, or more scope to merge bubbles) could offer support to isolated patients, but also isolated workers.

Above all, it is vital to recognize the specific contributions made by community health care professionals in navigating the multiple challenges of COVID-19 lockdowns. Rather than collapsing these into an undifferentiated category of ‘essential’ or ‘frontline’ worker, these distinct contributions must be acknowledged in accounts of the current pandemic, and be anticipated in future pandemic planning.

## Data Availability

The data underlying this study are not publicly available due to ethical and privacy restrictions.

## Acknowledgments

We would like to thank all the research participants who gave generously of their time and insights to help us understand their experiences of lockdown. Although they did not contribute to the writing of this article, Pounamu Jade Aikman, Naseem Jivraj, and Michael Roguski all made important contributions to data collection and preliminary analysis.

## References

Appleton, N. S., Long, N. J., Aikman, P. J., Davies, S. G., Deckert, A., Fehoko, E., Holroyd, E., Jivraj, N., Laws, M., Martin-Anatias, N., Roguski, M., Simpson, N., Sterling, R., Trnka, S., & Tunufa’i, L. (2020). (Alter)narratives of ‘winning’: supermarket and healthcare workers’ experiences of COVID-19 in Aotearoa New Zealand. SITES, 17(2), 51–77.

Bagnasco, A., Zanini, M., Hayter, M., Catania, G., & Sasso, L. (2020). COVID 19—A message from Italy to the global nursing community. Journal of Advanced Nursing, 76(9), 2212–2214.

Ballard, M., Bancroft, E., & Nesbit, J. et al. (2020). Prioritising the role of community health workers in the COVID-19 response. BMJ Global Health, 5, e002550.

Bear, L., Simpson, N., Bazambanza, C., Bowers, R. E., Kamal, A., Lohiya, A. G., Pearson, A., Vieira, J., Watt, C., & Wuerth, M. (2021). Social infrastructures for the post-Covid recovery in the UK. LSE. http://eprints.lse.ac.uk/111011/

Blanco-Donoso, L. M., Moreno-Jiménez, J., Gallego-Alberto, L., Amutio, A., Moreno-Jiménez, B., & Garrosa, E. (2021). Satisfied as professionals, but also exhausted and worried!! The role of job demands, resources and emotional experiences of Spanish nursing home workers during the COVID-19 pandemic. Health and Social Care in the Community, 00, 1–13. https://doi.org/10.1111/hsc.13422

Bowers, B., Pollock, K., Oldman, C., & Barclay, S. (2021). End-of-life care during COVID-19: opportunities and challenges for community nursing. British Journal of Community Nursing, 26(1), 44–46. https://doi.org/10.12968/bjcn.2021.26.1.44

Braun, V., & Clarke, V. (2019). Reflecting on reflexive thematic analysis, Qualitative Research in Sport. Exercise and Health, 11(4), 589–597, DOI: 10.1080/2159676X.2019.1628806

Brazeau, C., Schroeder, R., Rovi, S., & Boyd, L. (2010). Relationships between medical student burnout, empathy, and professionalism climate. Academic Medicine, 85(10), S33–S36. doi: 10.1097/ACM.0b013e3181ed4c

Brinkman, A. (2020). Coping with the COVID-19 virus. Kai Tiaki: Nursing New Zealand, 26(3), 25.

Brooks, S. K., Dunn, R., Amlôt, R., Rubin, G. J., & Greenberg, N. (2018). A systematic, thematic review of social and occupational factors associated with psychological outcomes in healthcare employees during an infectious disease outbreak. Journal of Occupational and Environmental Medicine, 60(3), 248–257. DOI: https://doi.org/10.1097/JOM.0000000000001235

Corcoran, L. (2021). Delivering wound care during the pandemic. British Journal of Community Nursing, 26(S6), S34–S37.

Cousins, S. (2020). New Zealand eliminates COVID-19. The Lancet, 395(10235), 1474.

De Kock, J. H., Latham, H. A., Leslie, S.J., Grindle, M., Munoz, S-A., Ellis, L., Polson, R. & O’Malley, C.M. (2021). A rapid review of the impact of COVID-19 on the mental health of healthcare workers: implications for supporting psychological well-being. BMC Public Health, 21, 104. https://doi.org/10.1186/s12889-020-10070-3

De los Santos, J.A.A., & Labrague, L. J. (2021). The impact of fear of COVID-19 on job stress, and turnover intentions of frontline nurses in the community: A cross-sectional study in the Philippines. Traumatology, 27(1), 52–59.

Franzosa, E., Gorbenko, K., Brody, A., Leff, B., Ritchie, C., Kinosian, B., Ornstein, K., & Federman, A. (2021). “At Home, with Care”: Lessons from New York City home-based primary care practices managing COVID-19. Journal of the American Geriatrics Society, 29(2), 300–306.

Fernandez, R., Lord, H., Halcomb, E., Moxham, L., Middleton, R., Alananzeh, A. & Ellwood, L. (2020). Implications for COVID-19: A systematic review of nurses’ experiences of working in acute care hospital settings during a respiratory pandemic. International Journal of Nursing Studies, 111, 10363. https://doi.org/10.1016/j.ijnurstu.2020.103637.

Green, J., Doyle, C., Hayes, S., Newnham, W., Hill, S., Zeller, I., Graffin, M., & Goddard, G. (2020). COVID-19 and district and community nursing. British Journal of Community Nursing, 25(5), 213. https://doi.org/10.12968/bjcn.2020.25.5.213

Green, J. (2021). RCN survey reveals ‘relentless pressures’ on district nurse services. Primary Health Care, 31(2), 11–13.

Halcomb, E., McInnes, S., Williams, A., Ashley, C., James, S., Fernandez, R., Stephen, C., & Calma, K. (2020). The experiences of primary healthcare nurses during the COVID-19 pandemic in Australia. Journal of Nursing Scholarship, 52(5), 553–563. https://doi.org/10.1111/jnu.12589

Holroyd, E. and McNaught, C. (2008), The SARS crisis: reflections of Hong Kong nurses. International Nursing Review, 55: 27–33. https://doi.org/10.1111/j.1466-7657.2007.00586.x

Houkamau, C. A., & Sibley, C. G. (2019). The role of culture and identity for economic values: a quantitative study of Māori attitudes. Journal of the Royal Society of New Zealand, 49(S1), 119–136. https://doi.org/10.1080/03036758.2019.1650782

Ives, J., Greenfield, S. & Parry, J.M. (2009). Healthcare workers’ attitudes to working during pandemic influenza: a qualitative study. BMC Public Health, 9, 56. https://doi.org/10.1186/1471-2458-9-56

Jackson, J., Posch, K., Bradford, B., Hobson, Z., Kyprianides, A., Yesberg, J. (2020). The lockdown and social norms: why the UK is complying by consent rather than compulsion. British Politics and Policy at LSE [blog], https://blogs.lse.ac.uk/politicsandpolicy/lockdown-social-norms/

Kelly, R, Wright-St Clair, V, Holroyd, E. (2020). Patients’ experiences of nurses’ heartfelt hospitality as caring: a qualitative approach. Journal of Clinical Nursing. 29 (11-12): 1903–1912. https://doi.org/10.1111/jocn.14701

Kvalsvig, A., & Baker, M. G. (2021). How Aotearoa New Zealand rapidly revised its Covid-19 response strategy: lessons for the next pandemic plan. Journal of the Royal Society of New Zealand, 51(S1), S143–S16. https://doi.org/10.1080/03036758.2021.1891943

Liamputtong, P. (2019). Qualitative research methodology and evidence-based practice in public health. In P. Liamputtong (Ed), Public health: local and global perspectives (2nd ed.). Cambridge University Press, Melbourne.

Liu, Q., Luo, D., Haase, J. E., Guo, Q., Wang, X. Q., Liu, S., Xia, L., Liu, Z., Yang, J., & Yang, B. X. (2020). The experiences of health-care providers during the COVID-19 crisis in China: a qualitative study. The Lancet Global Health, 8(6), e790–e798. ISSN 2214-109X, https://doi.org/10.1016/S2214-109X(20)30204-7.

Long, N. J. (2020). From social distancing to social containment: Reimagining sociality for the coronavirus pandemic. Medicine Anthropology Theory, 7(2), 247–260. https://doi.org/10.17157/mat.7.2.791

Long, N. J., Aikman, P. J., Appleton, N. S., Davies, S. G., Deckert, A., Holroyd, E., Jivraj, N., Laws, M., Simpson, N., Sterling, R., Trnka, S., & Tunufa’i, L. (2020). Living in bubbles during the coronavirus pandemic: Insights from new zealand http://eprints.lse.ac.uk/104421/

Long, N. J., Aikman, P. J., Appleton, N. S., Davies, S. G., Deckert, A., Fehoko, E., Holroyd, E., Jivraj, N., Laws, M., Martin-Anatias, N., Roguski, M., Simpson, N., Sterling, R., Trnka, S., & Tunufa’i, L. (forthcoming). On epidemiological consciousness and COVID-19: Envisioning vulnerability, hazard, and public health policy in Aotearoa New Zealand and the United Kingdom. In J. Robinson, S. Abram, & H. Lambert (Eds.), How to live through a pandemic. [ASA Volume; under review].

Lukewich, J., Poitras, M.-E., & Mathews, M. (2021). Unseen, unheard, undervalued: advancing research on registered nurses in primary care. Practice Nursing, 32(4), 158–162. https://doi.org/10.12968/pnur.2021.32.4.158

McClunie-Trust, P. (2021). Pandemic’s impact on the profession. Kai Tiaki: Nursing New Zealand, 27(3), 13–15.

Maben, J., & Bridges, J. (2020). Covid-19: Supporting nurses’ psychological and mental health. Journal of Clinical Nursing, 29(15-16), 2742–2750.

Major, C. (2008). Affect Work and Infected Bodies: Biosecurity in an Age of Emerging Infectious Disease. Environment and Planning A, 40(7), 1633–1646.

Marchesi, M. (2020). From sociality to social distancing: reversing values of solidarity in Italy. Social Anthropology, 28(2), 318–319.

New Zealand Government. (2020). Unite against COVID-19. Available at: https://covid19.govt.nz/

Nyashanu, M., Pfende, F., & Ekpenyong, M. S. (2020). Triggers of mental health problems among frontline healthcare workers during the COVID-19 pandemic in private care homes and domiciliary care agencies: Lived experiences of care workers in the Midlands region, UK. Health Soc Care Community, 00, 1–7. https://doi.org/10.1111/hsc.13204

Nyatanga, B. (2021). When death is part of us: Supporting community nurses during the COVID-19 pandemic. British Journal of Community Nursing, 26(3), 150–151.

Oldman, C. (2020). Exceptional community nursing care during the COVID-19 pandemic. Primary Health Care, 30(3), 5.

Penfold, J. (2020). Has the COVID-19 crisis changed community nursing in the UK forever? Primary Health Care, 30(3), 13–15.

Savitsky, B., Radomislensky, I., & Hendel, T. (2021). Nurses’ occupational satisfaction during Covid-19 pandemic. Applied Nursing Research, 59, 151416.

Shan, M., Peter, E., Killackey, T., & Maciver, J. (2021). The “nurse as hero” discourse in the COVID-19 pandemic: A post structural discourse analysis, International Journal of Nursing Studies, 117, 103887. https://doi.org/10.1016/j.ijnurstu.2021.103887.

Stanley, S., & Rawlinson, M. (2021). The only constant in life is change -- a case study of new working practices for podiatry and district nursing due to COVID-19. Diabetic Foot Journal, 24(1), 46–50.

Sumner, J., & Townsend-Rocchiccioli, J. (2003). Why are Nurses Leaving Nursing? Nursing Administration Quarterly, 27(2), 164–171. https://doi.org/10.1097/00006216-200304000-00010

Sun, Y., Song, H., Liu, H., Mao, F., Sun, X., & Cao, F. (2020). Occupational stress, mental health, and self-efficacy among community mental health workers: A cross-sectional study during COVID-19 pandemic. International Journal of Social Psychiatry. https://doi.org/10.1177/0020764020972131

Sun, S., Xie, Z., & Yu, K. Jiang, B (2021). COVID-19 and healthcare system in China: challenges and progression for a sustainable future. Global Health, 17(14). https://doi.org/10.1186/s12992-021-00665-9

Trnka, S., Long, N. J., Aikman, P. J., Appleton, N. S., Davies, S. G., Deckert, A., Fehoko, E., Holroyd, E., Jivraj, N., Laws, M., Martin-Anatias, N., Roguski, M., Simpson, N., Sterling, R., & Tunufa’i, L. (2021). Negotiating risks and responsibilities during lockdown: Ethical reasoning and affective experience in Aotearoa New Zealand. Journal of the Royal Society of New Zealand, 51(S1), S55–S74. https://doi.org/10.1080/03036758.2020.1865417

Trnka, S., & Trundle, C. (2017). Competing responsibilities: Reckoning personal responsibility, care for the other, and the social contract in contemporary life. In S. Trnka & C. Trundle (Eds.), Competing responsibilities: The politics and ethics of contemporary life (pp. 1–26). Duke University Press.

Trueland, J. (2020). Coronavirus in the community: the innovations helping nursing services keep up. Primary Health Care, 30(5), 6–8. https://doi.org/10.7748/phc.30.5.6.s2

World Health Organization. (2020). Coronavirus disease (COVID-19) outbreak: rights, roles and responsibilities of health workers, including key considerations for occupational safety and health. Available online at https://apps.who.int/iris/bitstream/handle/10665/331510/WHO-2019-nCov-HCWadvice-2020.2-eng.pdf

Wright St. Clair, V., & Seedhouse, D. (2005). The moral context of practice and professional relationships. In Whiteford, G. & Wright-St. Clair, V. (Eds.), Occupation and practice in context (pp. 16–33). Sydney, Australia: Elsevier.

Yi, X., Binte Jamil, N., Gaik, I. T. C., & Fee, L. S. (2020). Community nursing services during the COVID-19 pandemic: the Singapore experience. British Journal of Community Nursing, 25(8), 390–395.

Yıldırım, M., Arslan, G., & Özaslan, A. (2020). Perceived Risk and Mental Health Problems among Healthcare Professionals during COVID-19 Pandemic: Exploring the Mediating Effects of Resilience and Coronavirus Fear. International Journal of Mental Health Addiction, 16:1–11.

